# Gut microbiota features on nursing home admission are associated with subsequent acquisition of antibiotic resistant organism colonization

**DOI:** 10.1101/2020.04.25.20078428

**Authors:** Joyce Wang, Marco Cassone, Kristen Gibson, Bonnie Lansing, Lona Mody, Evan Snitkin, Krishna Rao

## Abstract

Nursing home (NH) patients often acquire colonization with antibiotic-resistant organisms (AROs). We show that patients exposed to broad-spectrum antibiotics during previous hospitalizations have elevated enterococcal relative abundances on NH admission and higher risk of subsequent ARO acquisition. Our findings suggest that interventions preventing ARO spread should extend beyond NH doors.

## Introduction

Antibiotic resistance is a major public health threat, particularly among vulnerable populations in healthcare settings [1]. Within the healthcare system, post-acute care settings such as nursing homes (NHs) have been recognized as an important reservoir for antibiotic resistant organisms (AROs) [2]. NH patients are often older adults with complicated medical conditions, and ARO acquisition is common in this patient population. In a recent study, one-third of patients uncolonized at the time of NH admission acquire an enteric ARO such as vancomycin-resistant *Enterococcus* (VRE) or resistant gram-negative bacteria (R-GNB) during their stay [3].

The role of intestinal microbiota in mediating the risk of ARO colonization has become increasingly appreciated in clinical settings where microbiota disruptions often precede ARO colonization [4]. While most research and intervention efforts have focused on improving appropriate antibiotic use in the NH setting, the impact of frequent antibiotic exposure during previous acute care hospitalization on ARO acquisition during NH stay has been mostly overlooked [3,5]. We hypothesized that due to the lasting effects of antibiotic exposure, antibiotic use at connected healthcare facilities could impact risk of ARO acquisition once in NHs. To test this hypothesis, we conducted a prospective, longitudinal cohort study to examine patient clinical factors and gut microbiota features at the time of NH admission and their associations with the risk of acquisition of VRE and/or R-GNB within 14 days of study enrollment.

## METHODS

### Study Population

From September 2016–August 2018, we enrolled patients from six NHs in southeast Michigan within 14 days of admission as part of a larger NIH-funded trial. Colonization status was determined by culture swabs collected from multiple body sites at enrollment, day 7, and day 14 using microbiological procedures described previously [3]. Our analysis focused on patients: 1) colonized with neither VRE nor R-GNB at the time of study enrollment, 2) had a perirectal swab (Copan ESwab™) collected at enrollment, and 3) had at least one follow-up visit with culture samples. Patient clinical data were collected at enrollment by trained research personnel for each patient using hospital transfer records and/or NH medical data. All subjects or their proxy provided a written informed consent. This study was approved by the University of Michigan Institutional Review Board.

## 16S rRNA Gene Sequencing and Analysis

Perirectal specimens underwent DNA extraction, 16S rRNA PCR-amplification, and MiSeq sequencing as previously described [6]. The resulting sequences were clustered into operational taxonomic units (OTUs) using *mothur* (see Supplementary Materials for detailed methods) [7]. OTUs typically associated with the skin microbiota at the genus level, including *Staphylococcus, Corynebacterium* and *Propionibacterium* were removed from downstream analyses.

## Statistical Analysis

All data analyses were performed in R (version 3.6.1). The primary outcome was the acquisition of VRE or R-GNB within 14 days of study enrollment. Exposures of interest included patient clinical factors and microbiota features at enrollment. Patient characteristics associated with outcome were determined by linear or logistic regression analyses. For microbiota features, we calculated the relative abundance of each OTU in each patient. As perirectal samples are low in biomass and rare OTUs may not be well-represented across samples, we conservatively restricted our analyses to only the 50 most abundant OTUs in this cohort. Linear regression analyses were used to examine associations between the relative abundance of each OTU and antibiotic exposure. Logistic regression analyses were used to identify associations between outcome and individual OTU, the fraction of anaerobes and butyrate-producers (see Supplementary Materials for details). Skewed data were log-transformed to achieve normal distribution. For each OTU associated with ARO acquisition with an unadjusted *P*-value of <.05, multivariable logistic regression analyses were used to identify patient factors that modified the effect of the OTU by >10%. To assess variables representing more global measures of microbiota state, we adapted a recently developed Microbiome Health Index (MHI) [8]. An MHI of 0 after log transformation indicates a balanced abundance between taxa associated with protection (*Bacteroidia* and *Clostridia*) and dysbiosis (*Gammaproteobacteria* and *Bacilli*). An MHI above 0 suggests higher abundances of protective taxa, and vice versa.

## RESULTS

### Exposure to antibiotics disruptive to the gut microbiota prior to NH admission is associated with ARO acquisition during NH stay

Of 243 patients enrolled in the study, 61 patients met our inclusion criteria: no VRE or R-GNB colonization on enrollment, a perirectal swab collected at enrollment and at least one follow-up within 14 days (Supplementary **Figure S1**). Most patients (73%) were enrolled within 7 days of NH admission (median, 6.0 days; range, 1 - 13 days).

Within 14 days of enrollment, 18 (29.5%) acquired AROs (3 VRE, 13 R-GNB, 2 both). In bivariable analyses, exposure to high-risk antibiotic classes with previously-demonstrated associations with gut microbiota disruption [9], including 3^rd^/4^th^-generation cephalosporins, quinolones, lincosamides, penicillin combinations, carbapenems and oral vancomycin prior to enrollment, was a significant risk factor associated with ARO acquisition (OR, 4.20; 95%CI, 1.08 -17.63). Notably, the majority of these high-risk antibiotics were initiated during hospitalization prior to admission to NH (N = 11/12), many for infection-related reasons including urinary tract infection, pneumonia and *Clostridium difficile* infection (Supplementary **Figure S2**).

## Microbiota features at enrollment differed between patients with and without ARO acquisition during NH stay

Following the observation that the receipt of high-risk antibiotics prior to NH admission was associated with risk for ARO acquisition; we next asked if this exposure had resulted in detectable disruption to the gut microbiota on admission to NHs. To assess this, we performed 16S sequencing on perirectal swabs taken from the 61 patients on enrollment. After removing skin-associated taxa that were not deemed as representative of the gut microbiota, 2326 OTUs were retained for final analysis. Considering only the 50 most abundant OTUs, we found that exposure to high-risk antibiotics prior to enrollment was associated with decreased relative abundance of several OTUs associated with the *Clostridia, Bacteroidia*, and *Actinobacteria* classes, and increased relative abundance of *Enterococcus* (**Figure 1A**).

**Figure 1.**
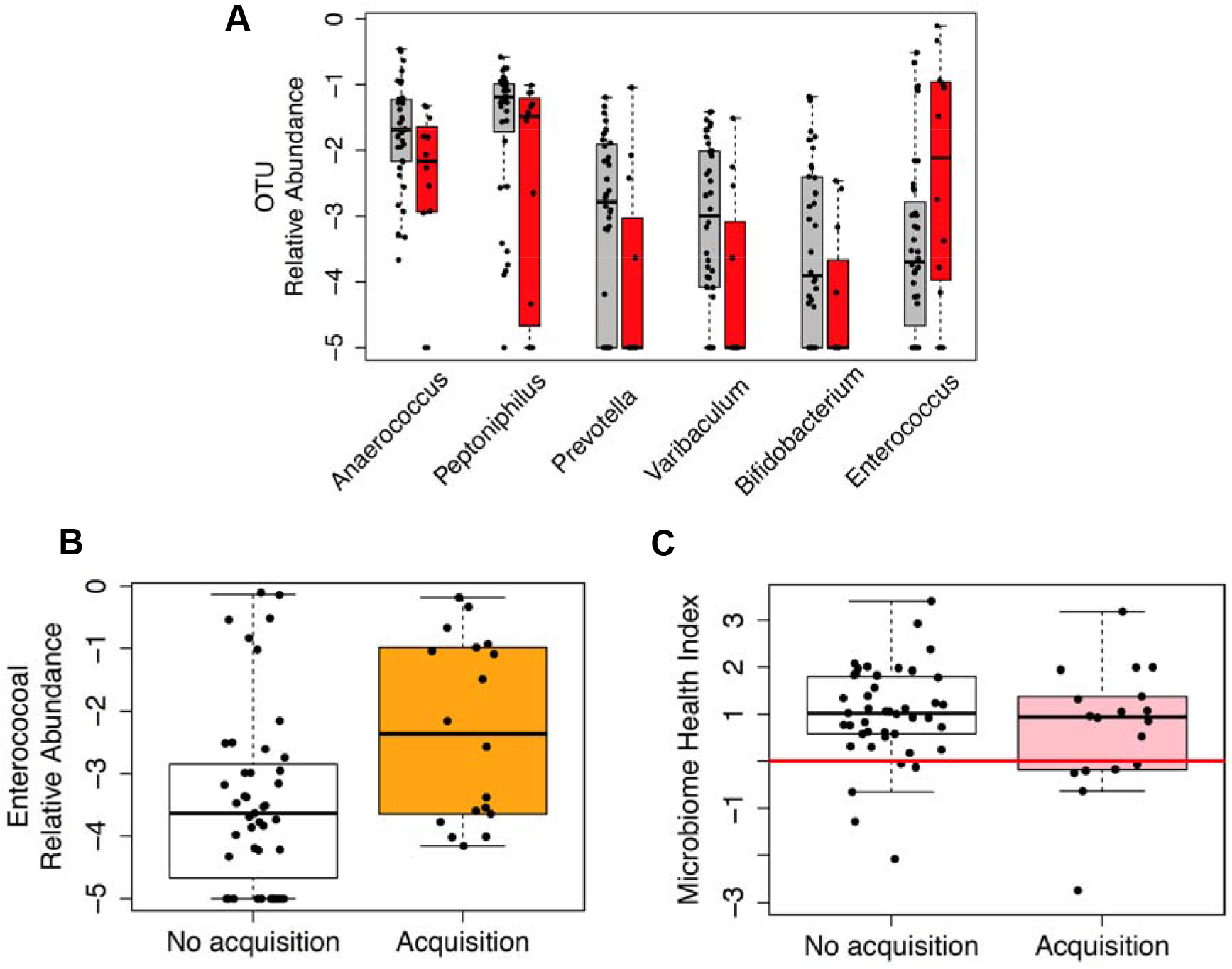
Microbiota features are associated with exposure to antibiotics during prior hospitalization and the risk of acquiring an enteric antibiotic-resistant organism (ARO) within 14 days. **A**. Relative abundances of operational taxonomic units (OTUs) significantly affected by prior exposure to high-risk antibiotics (P < .05). Without exposure: grey; with exposure: red. *Anaerococus* and *Peptoniphilus* belong to class *Clostridia, Prevotella* belongs to class *Bacteroidia, Varibaculum* and *Bifidobacterium* belong to class *Actinobacteria*, and *Enterococcus* belongs to class *Bacilli*. **B**. Relative abundances of *Enterococcus* in patients who did not acquire an ARO in 14 days (white) and those who did (yellow). **C**. Microbiome health index (MHI) in patients who did not acquire an ARO in 14 days (white) and those who did (pink). Red line indicates an MHI of 0. Median and interquartile range are shown in boxplots. The y-axes are presented on a base 10 logarithmic scale.

Next we looked directly at whether there were OTUs associated with risk for ARO acquisition, and found that patients who acquired an ARO during NH stay had significantly increased relative abundance of *Enterococcus* at the time of enrollment compared to those who did not acquire an ARO (log-transformed mean ±standard deviation [sd]: -2.31 ±1.46 vs -3.41 ±1.44, *P* =.012; **Figure 1B**). Incorporating clinical variables, including age, sex, race, body mass index, functional status, comorbidity scores, length of previous hospital stay, or urinary catheter use within the past 30 days in multivariable models did not significantly improve model fit, and the statistical significance of *Enterococcus* was retained, suggesting that altered *Enterococcus* abundance was an independent risk factor for ARO acquisition. Patients who acquired an ARO had lower relative abundances of anaerobes and butyrate producers at enrollment than those who did not, but the difference was not statistically significant. Lastly, examination of the microbiome health index (MHI) revealed that 6 out 18 (33.33%) of the patients who acquired an ARO had an MHI of < 0, which was significantly higher than the 5 out of 43 (11.63%) patients with no acquisition (chi-squared test *P* =.04) (**Figure 1C**). The association between an MHI of less than 0 and risk of acquisition remained significant after adjusting for hospital length of stay and exposure to urinary catheter within the past 30 days, both potential confounders that modified the effect of MHI by more than 10% (adjusted OR, 6.85; 95% CI, 1.46-39.24).

## DISCUSSION

With an aging population in the U.S., patients are more medically complex and likely to require post-acute care after hospitalization. ARO acquisition during NH stay not only negatively impacts patient outcome by increasing the risk of infection, but also contributes to the spread of resistance within NHs, between healthcare facilities, and the community [3]. By reviewing patient medical records, we found that recent exposure to high-risk antibiotics, mostly initiated during prior hospitalization, was associated with the risk of ARO acquisition. We further found that this association was mediated by microbiota disruption, as patients who subsequently acquired an ARO had detectable differences in their microbiota features at the time of NH admission.

Our finding that patients with elevated *Enterococcus* abundances were at a higher risk of enteric ARO acquisition was consistent with previous studies demonstrating that antibiotic therapy can promote high colonization density of VRE and R-GNB [4,10]. Of note, while *Enterococcus* was detected in all 18 patients who acquired an enteric ARO, only 3 acquired VRE, and the rest acquired either R-GNB (N = 13) alone or R-GNB with VRE (N = 2), suggesting that enterococcal expansion is a biomarker for susceptibility to enteric ARO colonization, instead of proliferation of pre-existing VRE colonization. Several patients with elevated *Enterococcus* abundances did not acquire ARO with 14 days, with these patients potentially representing an at-risk population that had a lower intensity of ARO exposure from other patients, healthcare workers, or the environment.

Our characterization of the gut microbiota of patients traversing the healthcare network demonstrates that exposure to high-risk antibiotics on NH admission influenced patient gut microbiota features and the risk of ARO acquisition during NH stay. Our study highlights the connectedness of the healthcare network and suggests the future study of coordinated infection prevention approaches between healthcare partners would be a fruitful endeavor [11].

## Data Availability

16S rRNA data will be released upon manuscript publication as a peer-reviewed article, or by Dec 31, 2020 whichever comes first. Selected metadata are available in the interim.

https://dataview.ncbi.nlm.nih.gov/object/PRJNA627982?reviewer=t17pps8bn6rrfiliapqnf6mgrj

## ACKNOWLEDGMENTS

We thank the patients and their families, and nursing homes who participated in this study; we thank members of the Mody laboratory for data collection and analysis, Drs. Christine Bassis and Jonathan Golob for technical and scientific discussions, and the Microbial Systems Molecular Biology Laboratory at the University of Michigan for performing 16S rRNA sequencing.

## FUNDING

This work was supported by the Centers for Disease Control and Prevention [Contract BAA 2016-N-17812 to E.S.S.], National Institutes of Health [grant number R01AG041780, K24AG050685 to LM], University of Michigan Host Microbiome Initiative Microbiome Explorers Program to E.S.S. the Canadian Institutes of Health Research fellowship [grant number 201711MFE-396343-165736 to JW] and the

Michigan Institute for Clinical and Health Research (MICHR) Postdoctoral Translational Scholars Program (PTSP) to JW.

## Notes

### Competing Interest Statement

The authors have declared no competing interest.

